# The Experience of Care Partners of Patients with Parkinsonism and Psychosis

**DOI:** 10.1101/2020.12.28.20248943

**Authors:** Sneha Mantri, Emily Klawson, Steven Albert, Robyn Rapoport, Chelle Precht, Sarah Glancey, Margaret Daeschler, Eugenia Mamikonyan, Catherine M. Kopil, Connie Marras, Lana M. Chahine

## Abstract

**Background:** Parkinson’s disease psychosis (PDP) has a major impact on quality of life and care partner burden; however, little is known about the lived experiences of care partners in managing PDP.

**Objective:** To understand how care partners of individuals with PDP experience their role and articulate their needs related to psychosis.

**Methods:** This was a qualitative study of semi-structured telephone interviews. Recruitment was conducted online via the clinical study matching tool, Fox Trial Finder; study activities took place remotely via telephone interviews. Transcripts of the phone interviews were analyzed by grounded theory methods, and a codebook of key themes that emerged from the analysis was developed.

**Results:** Nine care partners (all female) were interviewed. Discussion topics in the codebook included (1) care partner burden and guilt; (2) communication with medical professionals; (3) coping strategies; (4) emotional reactions of the care partner to psychosis; (5) sources of knowledge about PD psychosis; (6) attitudes towards medications for PDP; (7) strategies to care for loved ones with psychosis; (8) psychosis triggers.

**Conclusions:** This qualitative analysis uncovers important aspects of the care partner experience, including challenges in navigating the medical system and communicating with professionals. Providers treating patients with PDP should be aware of these constraints and provide added support for strained care partners.

## INTRODUCTION

Psychosis is common in advanced Parkinson’s disease (PD), with a cumulative prevalence of over 80% [1]. Individuals with PD who suffer from psychosis have decreased quality of life and increased health resource utilization [2]; care partners of these individuals also have higher levels of care partner burden [3]. Additionally, care partners provide a large amount of unpaid care for loved ones with PD psychosis (PDP), leading to higher indirect costs (e.g., disability and medically-related absenteeism-related to caring for a loved one) and higher cumulative income loss over time [4]. Thus, minimizing care partner strain may have important impacts on both psychosocial and economic burden.

Nevertheless, despite increasing recognition of and treatment strategies for PDP in recent years, little is known about the lived experience of care partners navigating this challenging phase of illness. Prior qualitative research has suggested that care partners may have difficulties communicating with medical providers about both motor and non-motor aspects of disease [5,6]; understanding these difficulties would be valuable to design strategies to improve caregiver-physician communication In this study, we examined semi-structured interviews with care partners to discern common struggles around communication with providers, navigating available community resources, and the economic and psychosocial impact of caring for a loved one with PDP.

## MATERIALS AND METHODS

The online clinical study matching tool Fox Trial Finder (FTF) (https://foxtrialfinder.michaeljfox.org) was used to identify care partners for participation in this study. As previously described [7], FTF is a database of PD-related research volunteers. Self-identified care partners enrolled in FTF were sent an email invitation to participate. The email invitation included a link to a survey which assessed basic eligibility criteria for inclusion in this study: English-speaking, age 18 or older, resident of the USA, and access to a telephone for interview. The initial criteria included only care partners living with PD patients, but these were later modified to allow care partners not living with the patient to also participate. There were 193 respondents to the invitation, and 86 met inclusion criteria. A convenience sample of consecutive respondents was contacted for participation. Verbal informed consent was obtained over the telephone. A screening interview was then performed. Eligible subjects were administrated the Symptoms for Parkinson’s Disease Psychosis (SAPS-PD) scale over the telephone, with the goal of obtaining preliminary information on the burden of psychosis the recipient of care has. The SAPS-PD [8] is a 9-item scale interviewer-administered scale derived from the Scales of Assessment of Positive Symptoms (SAPS) that asks about positive symptoms of psychosis.

The study was conducted in two phases (Figure 1). For the first phase of a study, the goal was to recruit 20 care partners to participate in prompt-driven online journaling activities. The objective of this phase was to collect preliminary, pilot data to inform next phases. A convenience sample of 47 consecutive respondents were contacted until 20 had been recruited for participation in the online journaling. This consisted of a series of activities, grouped into three sessions, in which subjects responded to prompts, pictures and graphics with free-text responses. Each session was estimated to take 30 minutes. Participants were asked to complete one session a day over three days but had up to one week to complete all the sections. Responses were informally analyzed by the study team to assess components of psychosis most important to care partners and to inform development of a semi-structured discussion guide to be used during subsequent telephone interviews.

**Figure 1.**
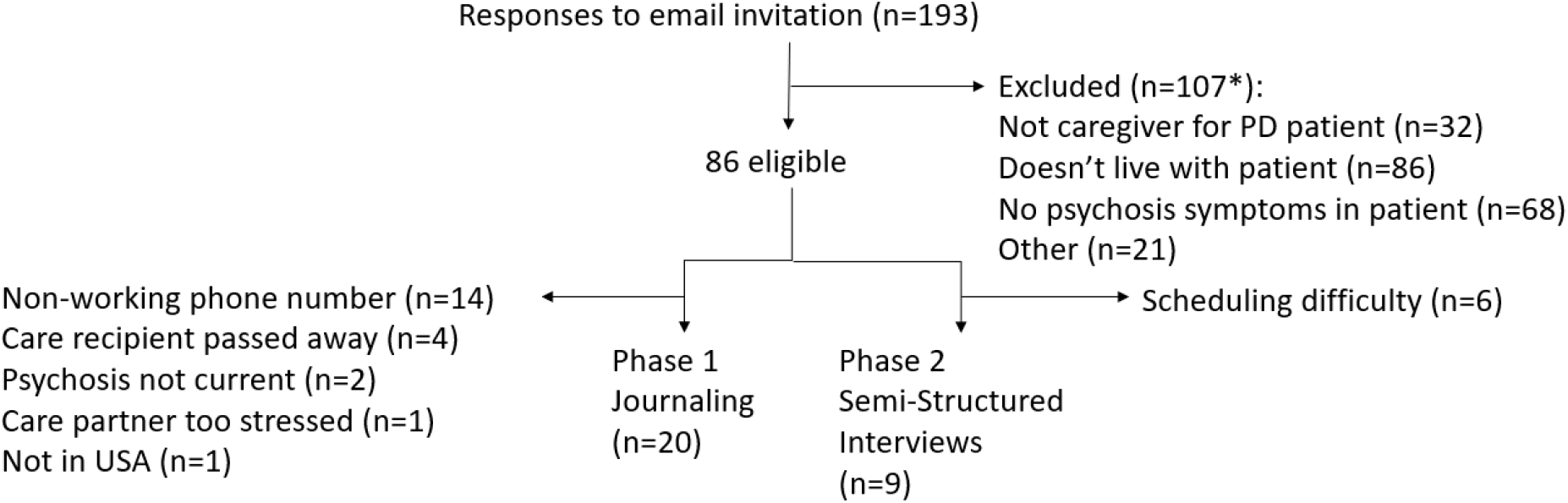
Flow Chart of Participation

Following this, a sample of additional care partners was recruited to participate in telephone interviews. This sample was also drawn from respondents to the invitation on FTF. A discussion guide for a one-hour semi-structured telephone interview was developed. Interview topics, drafted based on the results of online journaling as well as expert opinion of neurologists with subspecialty expertise in movement disorders (authors SM, CM, LMC). Foci of discussion included duration and evolution of care needs, communication strategies with health care providers, and coping mechanisms employed by the care partners. Care partners were also asked to provide descriptions of psychotic episodes, including triggers and responses. The discussion guide was administered by a consultant trained in qualitative research methods (author CP).

Qualitative analysis of interview transcripts was performed by researchers trained in qualitative research methods (authors SM, SA, EK). NVivo 12 Pro was used to develop a codebook of common themes. Themes were refined by repeated, iterative discussion between researchers [9] until a single standardized codebook was developed. Based on the structure of the discussion guide, themes were organized into three major domains of (A) describing psychosis; (B) care partners’ physical and emotional well-being; and (C) interfacing with the health care system.

This study was performed in accordance with the Declaration of Helsinki. This study is approved by the New England Institutional Review Board, and informed consent is obtained from each participant at enrollment.

## RESULTS

The target sample at onset of recruitment was 15 individuals. However, due to challenges in recruitment, largely related to scheduling difficulties of participants within the pre-specified time frame for data collection, eventually, 9 telephone interviews were conducted. Participant characteristics and burden of psychosis in the care recipients are shown in Table 1. Mean age of care partners was 61.4 years (standard deviation 8.6 years), all female. Seven were spouses of the care recipient, 1 was a sibling, and 1 a child. Duration of parkinsonism in the care recipient ranged from 0.5-18.5 years.

**Table 1.** Demographic characteristics of Telephone Interview Participants.

| Care partner Characteristics |  |  | Person with PDP Characteristics |  |  |  |  |  |  |  |  |  |
| --- | --- | --- | --- | --- | --- | --- | --- | --- | --- | --- | --- | --- |
| ID | Age | Relation to care recipient | PD Duration (years) | Auditory Hallucinations | Voices Conversing | Somatic or tactile hallucinations | Visual hallucinations | Global rating severity of hallucinations | Persecutory delusions | Delusions of jealousy | Delusions of reference | Global rating severity of delusions |
| 1 | 72 | Spouse | 8 | 0 | 0 | 0 | 2 | 2 | 0 | 0 | 0 | 0 |
| 2 | 63 | Spouse | 5 | 0 | 0 | 2 | 4 | 2 | 0 | 0 | 0 | 0 |
| 3 | 70 | Spouse | 12 | 0 | 0 | 4 | 0 | 3 | 5 | 3 | 0 | 2 |
| 4 | 60 | Spouse | 8 | 0 | 0 | 0 | 3 | 3 | 0 | 0 | 2 | 2 |
| 5 | 54 | Sibling | 0.5 | 2 | 0 | 2 | 2 | 2 | 3 | 3 | 0 | 3 |
| 6 | 65 | Child | 18.5 | 0 | 0 | 0 | 3 | 2 | 0 | 0 | 0 | 0 |
| 7 | 53 | Spouse | 12 | 0 | 0 | 2 | 2 | 2 | 0 | 2 | 2 | 2 |
| 8 | 69 | Spouse | 17 | 2 | 0 | 2 | 2 | 2 | 2 | 0 | 0 | 2 |
| 9 | 47 | Spouse | 7 | 0 | 0 | 2 | 2 | 3 | 0 | 0 | 2 | 2 |

Care partner-reported psychosis symptoms in the care recipients, as ascertained with the Symptoms for Parkinson's Disease Psychosis (SAPS-PD) are also shown. SAPS-PD 0-5: 0=None; 1=Questionable; 2=Mild
3=Moderate; 4=Marked; 5=Severe

A detailed codebook can be found in the online Supplement. Topics of discussion included (1) care partner burden and guilt; (2) communication with medical professionals; (3) coping strategies; (4) emotional reactions of the care partner to psychosis; (5) sources of knowledge about PD psychosis; (6) attitudes towards medications for PD psychosis; (7) strategies to care for loved ones with psychosis; (8) psychosis triggers. In reviewing these topics of discussion, we arrived at three overarching domains of (A) description and characteristics of psychosis; (B) the impact of psychosis on care partners; (C) challenges faced by care partners in navigating the medical system and advocating for their loved one.

### A. Description and characteristics of psychosis

Many care partners were able to identify clear triggers for the care-recipient’s psychosis. Common triggers included environmental cues, time of day (e.g. “A lot of times he’ll be a lot worse in the evenings”) and wearing off of medications (e.g. “He has his off periods. You know what off periods are. It’s definitely worse when he is in those, with everything. With paranoia, yes. With everything, when he has an off period. It’s all worse with that.”) Some episodes seemed to have multiple triggers; one care partner provided a detailed description of a chaotic series of episodes she believed to be triggered by a combination of medication changes, medical intervention, and a new living situation:

*“His, and this is what I think triggered this because he’s really been pretty good, his neurologist, movement disorder specialist decided to wean him off of quetiapine, Seroquel because he has a slow heart rate and he had been collapsing some. She felt that he should be weaned off of that medication because his EKG showed that his heart rate was slow. What happened, I did it exactly. We went through several weeks and as soon as we got down to 12 and a half milligrams, had that for two days. I’m going to describe it as, shut down. Luckily I have a friend who’s a retired nurse, and she said, he has shut down. I called his primary and they said take him to the ER. The second day, which was the first full day he was in the ER, again I know what triggered it. They were doing an ultrasound to see if his carotid artery were plugged, they were doing all these, trying to figure out what was happening and he for whatever reason, went into a full-blown psychotic, wild, at least verbally, violent episode, which he had done in the Galapagos. The attending physician came, obviously, they called him, and he said he needs that quetiapine, and I said give it to him. He was better in the hospital then. They kept him awake. Then a week ago yesterday, Monday, he went to the skilled nursing facility and he was doing pretty well. I mean I was with him, he was tired, and of course, many people in the skilled nursing facility are elderly and hard of hearing. The CNA came in, happened to be a man, deep voice, loud voice and it triggered a psychotic. Again the similar kind of episode that it’s hard to describe. He rambles, there are terrorists, he’s all over the place but he’s a very mild-mannered person but he’s wild anyway. I asked the CNA to leave, I said I will deal with this, and I did*.*” (Care partner 3, care partner for 12 years, SAPS-PD of care recipient 17)*.

### B. The impact of psychosis on care partners

Most of the care partners reported providing a significant amount of unpaid care, from managing medications to attending doctors’ appointments, which prevented them from pursuing other commitments. One care partner stated “I did give up positions on two boards because I felt the stress of being overcommitted. Whereas, before in my life, I could do multiple things and balance them all.” Another denoted her caregiving responsibilities in explicitly work-based terms: “18.5 hour shifts straight multiple times a week.” For the majority of respondents (8 of 9), the subject was the sole care partner for the patient; the 9th care partner, a daughter, shared caregiving responsibility with her sisters, but with significant disruption to her own life: “I drop[ped] out of school and … come here for three weeks out of each month then new people can take the other week, so I can go home….This will be my eighth summer.” Care partners did not distinguish between the impact of providing psychosis-related care and the impact of providing care in general.

Care partners reported a variety of negative emotions around their caregiving responsibilities and changed relationships. One stated “after it’s gone on maybe an hour, I’m tired and I lose my temper and I go through, in more of a parental voice.” Another acknowledged that she was “in total denial,” and a third reported suicidal ideation, including “two or three occasions when I had to stop myself from pulling in front of” oncoming traffic. This care partner reported that she was hesitant to report these feelings because of fear that “the bad agency people [would] use that against me.” Other care partners, however, did acknowledge benefit from speaking with support groups, therapists, social workers, or physicians. Other important coping strategies included mindfulness meditation or other solitary activities such as listening to podcasts and reading books.

### C. Navigating the medical system and advocating for the PD care recipient

All care partners reported significant challenges navigating the medical system. Seven of nine care partners reported that the neurologist or movement disorders specialist managing Parkinson’s symptoms did not routinely ask about psychosis. Of the two who did obtain information about psychosis from a physician, both described being asked “a list of questions” or “he just ran down the examples. He didn’t really go into it too much.” Alternate sources of knowledge about psychosis included informal networks (e.g. “I talked to a friend about it and her mother had Parkinson’s”), support groups and workshops by national organizations, and television commercials or “doctor Google.” Care partners also expressed concern about wanting to shield the care recipient or themselves from embarrassment by openly discussing psychosis in the clinical visit, instead resorting to sending email or electronic medical record portal messages, phone calls, or passing notes through nurses or receptionists.

Care partners expressed ambivalence regarding the use of medications to treat PD psychosis. Concerns included fear of medication interactions: “It all depends on the side effects and whether they all play well together…. We’re walking a real fine line with the way all these are balanced together.” All care partners felt that the threshold to start medications for psychosis depended on when a patient was felt to be a danger to themselves or others. Of the three care partners whose PD partners had started antipsychotic medications, all were on quetiapine. Care partners reported feeling satisfied with responsiveness of psychosis to this medication; one likened an attempted taper (quoted above) as akin to breaking an addiction.

## DISCUSSION

This qualitative analysis of the experience of care partners of people with PD psychosis uncovers important dimensions of caring for an individual with an advanced neurodegenerative condition. Triggers of psychosis identified by care partners were consistent with triggers taught to medical professionals [2], including changes in environment or routine, sundowning, and the impact of medications on psychotic symptoms. Importantly, care partners reported relying on informal networks for knowledge and anticipatory guidance, and those who did discuss psychosis with their partner’s physician rarely did so in the presence of the patient. Additionally, the impact of psychosis on care partners’ emotional state should not be dismissed. All care partners experienced significant disruption to their own lives in caring for their partner, several reported feelings of isolation and depression, and one revealed thoughts of suicide. All commented on the importance of anticipatory guidance from the medical care team as their care recipient’s disease progressed. Surprisingly, the majority of care partners reported that they were never routinely asked about psychosis by their care recipient’s neurologist, despite the fact that psychiatric symptom assessment (including psychosis) is considered standard of care for people with PD [10]. This underscores an important practice gap that warrants further attention.

While care partner burden and care partner burnout is increasingly recognized to contribute to poor outcomes in PD [3,11,12], a qualitative interview captures the severity of care partner burden in a much richer way than a Likert-scale questionnaire. For instance, the extended quotation from care partner 3’s interview mirrors what Arthur Frank, a medical sociologist, terms the chaos narrative [13]. In the chaos narrative, the storyteller (here, the care partner) struggles to provide a coherent, linear plot; rhetorical structures may be repeated, settings may shift abruptly, and time may shrink or dilate without warning. This style produces in the reader a sensation of anxiety, akin to that felt by the care partner during the psychotic episode itself. Several of the care partners interviewed emphasized the need for routine and structure in mitigating psychosis symptoms. However, the chaotic lack of structure caused by psychosis, particularly psychosis without an apparent trigger, led to deep emotional distress among care partners, including reports of suicidal ideation. Understanding the dynamics of the chaos narrative may help clinicians comprehend the struggles of care partners at this challenging stage of the disease.

To our knowledge, this is the first such report of qualitative interviews of care partners dealing with PDP. The themes uncovered by our analysis are consistent with other qualitative analyses of PD care partner experience [14–16] and the experience of care partners of those with other causes of psychosis [17], including the challenges of self-care while caregiving and the need for better care partner support.

Noted strengths of the current study are detailed verbatim transcripts by highly articulate care partners, which allowed for development of a nuanced codebook. Nevertheless, some important caveats should be kept in mind. As with all qualitative studies, the number of interviews was relatively small, favoring depth of analysis over breadth. All the care partners were female and highly educated; their experiences may not necessarily reflect the challenges faced by care partners of other sociodemographic groups. In particular, quantitative data suggests that male and female care partners experience their role differently [18]; we were unable to verify this in our interviews as only female care partners volunteered to be interviewed.

Further, we did not have access to medical records to verify diagnoses, disease durations, or psychosis duration reported by care partners. One care partner reported that her care recipient had been diagnosed with PD just 6 months before; early development of psychosis in this individual might suggest an alternate diagnosis, such as dementia with Lewy bodies, and the different prognosis of this disease may warrant more intensive support than typical for idiopathic PD. Additionally, there was wide variation in duration of caregiving (range 6 months to 18.5 years), and care partner experience may vary according to how long they have been involved with their care recipient; this could be an interesting area of future study. Nevertheless, core themes such as the need for better care partner education and anticipatory guidance from physicians, are likely readily translatable to a variety of settings and can serve as a needs-assessment for the development of educational materials for patients and care partners around PDP.

## Data Availability

Data cannot be shared publicly because they consist of potentially identifiable audio recordings. This requirement is set by the New England IRB who approved the study. Data are available from the Michael J. Fox Foundation (contact: Bernadette Siddiqui) for researchers who meet the criteria for access to confidential data

## ACKNOWLEDGEMENTS

We would like to thank volunteers on The Michael J. Fox Foundation’s Fox Trial Finder, an online clinical trial matching tool for Parkinson’s research (www.michaeljfox.org/trial-finder).

## AUTHOR CONTRIBUTIONS

Conceptualization: CM, LC

Data Curation: EK, SA, EM

Formal Analysis: SM, EK, SA

Funding Acquisition: LC

Investigation: CP

Methodology: SM, LC

Project Administration: RR, CP, SG

Resources: MD, CK

Software: SM, EK

Supervision: CM, LC

Validation: SM, EK, SA

Visualization: SM, EK, SA

Writing Original Draft: SM

Writing/Review and Editing: EK, SA, RR, CP, SG, MD, EM, CK, CM, LC

## SUPPPORTING INFORMATION

**S1. Table. Codebook of Discussion Topics, Sub-codes, Definitions, and Representative Quotations**. Codes were defined by grounded theory methods and refined through iterative discussion among the research team.

