## Supplementary material for "The Experience of Care Partners of Patients with Parkinsonism and Psychosis": Online Supplement

**ONLINE SUPPLEMENT: CODEBOOK**

| **CODE NAME** | **DEFINITION** | **EXAMPLE** |
| --- | --- | --- |
| **1. BURDEN** | Explanation of the amount of time, energy, work, responsibilities, etc that the caregivers are providing for their family member with PD. | TI001: “I know that the caregiving experience has affected me. I did give up positions on two boards because I felt the stress of being overcommitted. Whereas, before in my life, I could do multiple things and balance them all.”  TI003: “I’m not a martyr, that isn’t it at all, but so far it’s been pretty good. Now, I will say that this psychosis in the middle of the night is really wearing, because then I don’t get enough sleep.”  TI006: “I take care of her 24/7, it amounts to about three weeks for every month for nine months of the year. We share the duties with some of my sisters. She pretty much can't do anything for herself”  TI008: “I was doing 18 and a half hour shifts straight multiple times a week. I was so exhausted I couldn't even think, much less stay awake and take care of him. He cannot be unattended at all.”  TI009: “How many hours per week do you spend with your husband, in care of his PD? TI009: We're together 24/7.” |
| 1a. BURDEN_GUILT | Mention of need to protect family member with PD from feeling guilt or negative feelings | TI001: “I try to talk about it with him, so he doesn’t feel guilt that he’s ruining my life or that’s keeping me from doing something that I would want to do.”  TI003: “I have explained to him, if I’m extremely tired in the morning, what happened at night. Because he doesn't remember it. Yes, he doesn’t remember. I will explain that, “This is what happened, this was Parkinson’s.” Because I don’t want him to feel guilty. He has basically no control over that.”  TI004: “If the appointment was somehow compartmentalized and they saw me separately from (my husband), because I really don’t want to hurt (my husband’s) feelings and say, “Boy honey, I think you’re a real loon part of the time.” It just makes him feel so bad.” |
| 1b. BURDEN_ALONE | Caregiver is sole caretaker for loved one with PD | TI002: “Are you the only person who provides care for your husband, or are there others that help? TI002: No, I’m the only one.”  TI007: “Are you the only person who provides care for your husband or are there others? TI007: It’s just me.” |
| **2. COMMUNICATION** | Caregiver communication with the physician or medical staff caring for their family member with PD. |  |
| 2a. COMM_CAREMENT | Caregiver brings up psychosis symptoms at physician’s office | TI008: “Do you ever talk with your doctor about your husband’s psychosis symptoms and how they affect you? TI008: Yes. About all of his Parkinson’s and how it affects me. I don't think the psychosis is worse than just the disease, the toll it takes on my life.”  TI009: “How long have the symptoms of psychosis been going on before your doctor asked about them for the first time? TI009: I don't know, six months maybe. A year. It wasn't something that got brought up. We maybe brought it up the first time.” |
| 2b. COMM_NOCAREMENT | Caregiver does not bring up psychosis symptoms at physician’s office | TI001: “Have you ever brought it up to them? TI001: No, because my husband is so open with me about things.”  TI006: “Do you ever talk with your doctor about how the psychosis symptoms affect you? TI006: No. Chelle: (27:52) Why not? TI006: I never did. Chelle: (27:56) Why? TI006: It didn’t bother me.” |
| 2c. COMM_MDMENT | Physician asks about psychosis symptoms at appointment | TI002: “What have you observed as a caregiver that the doctor might not know about your husband’s psychosis symptoms? TI002: He took an hour to basically talk with us. I think that he knows pretty much what's going on.”  TI004: “Try to think back, did you all bring it up first, you, or your husband, or did the neurologist bring it up first? TI004: I think the neurologist brought it up first.” |
| 2d. COMM_NOMDMENT | Physician does not ask about psychosis symptoms at appointment | TI007: “What about your husband’s movement specialist? Do they ever ask you about how it’s impacting you? TI007: No they do not.”  TI009: “Anyone ever ask about those? TI009: No. That's what I said. That's why we were so unaware of it because it had never been brought up in any of our appointments.” |
| 2e. COMM_METHOD | Method of communication with physician or doctor’s office | 1. E-mail:    a. TI001: “I do email the doctor. I do handle the communication for my husband between him and with that doctor.”    b. TI003: “I haven’t brought it up within an appointment, I have MyChart-ed, basically an email, and said, “I’m seeing this.” 2. List/writing:    a. TI001: “We usually come with a big list of things that we’ve noticed.”    b. TI008: “Then, as I said, he will, quite often, give us some very concrete suggestions and type them up. We bring them home and we put them in a little book for everybody to read. They sign and initial how they read it.” 3. Talking in-person:    a. TI008: “We talk about a lot of these issues. We also talk about how we try to address him, and then he’ll write up something that I can take home so that the other caregivers can help approach him differently to deal with whatever the issue is.” 4. Talking on phone/call:    a. TI009: “I have called and spoken with her nurses, and then she’ll get back to me. When something major happens, that he has seen stuff or done stuff, I just call and let them know…” 5. Passing notes:    a. TI004: “I know a couple times it’s like I’ve passed notes through the receptionist to say, “Here, have the doctor read this before we get in there.” |
| 2f. COMM_POS | Note a positive interaction or communication with physician | TI005: “But with my sister’s primary care, she listens to everything I say. She was the one that when I initially told her, she started the ball rolling. She did an assessment and that initially led to the neurologist diagnosing her with Parkinson’s once she was in the hospital. After all this had happened. Yes, I definitely felt heard now.”  TI008: “He's an old fashioned guy. He spends an hour with you. He sees my husband when he’s on and when he’s off. He talks to me to make sure I'm doing okay. We can call him on Sunday afternoon to fill up his psychotic meds and he’ll talk to him. He's just wonderful.” |
| 2g. COMM_NEG | Note of negative of unsatisfied interaction or communication with physician | TI004: “I actually think a lot of his symptoms are worse than what the doctors are giving him credit for if that makes sense. Chelle: (33:47) Yes, tell me why you think that. TI004: Because they’re not spending a whole lot of time with him, especially his neurologist.”  TI007: “I do think it maybe should be something that doctors do ask about. A lot of times the doctors don’t want to talk about it either.” |
| **3. COPING** | Coping strategies used by the caregiver to cope with stresses of being a caregiver/of PD. |  |
| 3a. COPING_ACTIVITY | Caregiver mentions activities as coping method | TI006: “I listen to podcasts, I read books, I watch TV on my iPad. I dream about buying a house and I keep looking at houses, which I'm never going to buy a house back here, but anyway that's I use myself.”  TI008: “What other methods are you using to help you cope? TI008: The one that works the best I haven't been doing lately. It's mindfulness meditation.” |
| 3b. COPING_ENTITY | Referring to PD as a separate entity or personified, distinct from the caregiver’s family member. | TI003: “What I have done with my husband is differentiated between my husband and Parkinson’s. I’ve personified Parkinson’s.”  TI008: “They try to remind me that's not your husband, that's the Parkinson’s. Actually the people in the house do that too. When they can do that, that usually helps me.” |
| 3c. COPING_FAMFRIEND | Caregiver talks with friends or family members as a coping method | TI004: “How about you? How do you take care of yourself? TI004: Spend a lot of time talking to the dog.”  TI004: “Yes. I vent to my older daughter a lot too.”  TI009: “Do you ever discuss how PD is affecting you, in general? TI009: Not with a specialist, no, just more with like family and friends.” |
| 3d. COPING_GROUP | Caregiver support group or other supportive group | TI002: “I have started going to a PD care partners meeting. We have a wonderful PD community here in Santa Cruz, and they have these care partner meetings where people meet up without their spouses and talk about these sorts of things just to say, “Oh yes, my husband does that, too.””  TI003: “I’m in a Spanish group, and what’s lovely is they listen if I want to talk, but we don’t necessarily talk about Parkinson’s.”  TI004: “I’ve been playing around with a couple Facebook groups that are... There are two of them that I’ve been a little more active with recently. One is a support group for caregivers of Parkinson's patients and the other one is a support group for caregivers, it’s for the wives of husbands who have Parkinson's.” |
| 3e. COPING_MED | Prescription of or use of medications for caregiver to cope | TI001: “I said to my doctor, I sought him, and I said, “You know, I think that antidepressant would be a good thing.” We talked about it and I said, “I’m not sure if this would be a long-term thing or if I’ll need to be taking for a long while.” He told me to expect that I might be on them for a while.” |
| 3f. COPING_NONE | Caregiver has not gone to support groups, professionals, etc for coping | TI004: “Have you ever been to a support group to help with psychosis symptoms for you? TI004: No I have not. Chelle: (42:34) Therapist, counselor, psychologist, psychiatrist? None of those? TI004: No.”  TI005: “What, if anything, are you doing for yourself to help you cope with your sister’s psychosis symptoms? TI005: Nothing. Chelle: (37:37) Have you ever been to a support group? TI005: (Laughs). No, I don’t like support groups.” |
| 3g. COPING_PROF | Caregiver seeking or considering seeking professional help, such as a therapist | TI002: “Do you think you'll ever go further than the caregiver meetings at this point? Thinking specifically about psychosis. TI002: Yes. I've been playing with the idea of having a therapist that I go to, just somebody to talk to one on one.”  TI003: “The social worker helped me with that. She said, “Yes, you have an umbrella of all of these responsibilities that probably will need to be done, but you don’t have to do them all right now. Choose what you think needs to be done right now.” That’s my long-winded comment as to I have talked to the social worker.” |
| 3h. COPING_RES | Caregiver describes resilience or acceptance in face of caring for loved one with PD | TI001: “…clearing my schedule a little bit so that I wasn’t so stressed was important. I’m proactive on my own behalf.”  TI002: “We’re sucking up and doing it. I don't think either of us really likes it. It's like, ‘Okay, we've got to. This is what needs to happen in order to mitigate the circumstances of this particular disease.’” |
| **4. EMOTION** | Descriptions of how PD/PD psychosis affects the emotions of or emotional reactions of the caregiver. | TI001: “I also said, I was feeling blue for a long time and I said to my doctor, I sought him, and I said, “You know, I think that antidepressant would be a good thing.””  TI002: “He said, “Well, there's things that I know aren't real but I see them.” Then I'm lie, “Was surprised,” because he hadn't told me about that.”  TI003: “I think after it’s gone on maybe an hour, I’m tired and I lose my temper and I go through, in more of a parental voice, “I am not lying to you, I will never lie to you, I don’t cheat on you, yadda, yadda, yadda.””  TI005: “I guess because I’m more worried about her than I am me.”  TI006: “The only thing that she ever did that really got to me was the crying that you could not console her. That was very upsetting to me, because I couldn't console her”  TI007: “And yes, he did tell me about it just because it scared the crap out of me.”  TI008: “He can't even acknowledge that you're there. That's devastating.”  TI008: “I just am in total denial. I cannot make myself say, “That's Parkinson’s, that's not your husband. Your husband is still the loving, kind person you married.”  TI009: “It was minus 30 here and he was going to go. He said, “I figured if I didn't know where I was going, I would just get out and walk.” It's stuff like that that scares me.” |
| **5. KNOWLEDGE** | Caregiver’s first source of or scope of knowledge about PD/PD psychosis. |  |
| 5a. KNOW_FAMFRIEND | Information from a family member or friend | TI002: “I talked to a friend about it and her mother had Parkinson’s.”  TI009: “Not really. My brother-in-law has just sent me a bunch of information on support groups for just the psychosis. I have that information, I just haven't looked into it more.” |
| 5b. KNOW_GROUP | Information from group (caregiver support group, etc) | TI001: “Is that where you first learned about psychosis, PD psychosis? Or was there some other avenue? TI001: I did hear about it before in the caregiver’s group first.”  TI008: “As I said, I've taken dementia classes offered by the Alzheimer’s Association and that I found out about once a year there tends to be a day long conference on Parkinson’s disease.” |
| 5c. KNOW_MD | Information about psychosis from physician | TI002: “When he was asking the questions did he tell you a definition of, “Here’s what a delusion is, here's what a hallucination is, here's paranoia,” or did he just run down the examples like I did? TI002: He just ran down the examples. He didn't really go into it too much, but we’re both college educated, we pretty much know that that's an issue.”  TI005: “Her PCP called me and asked me a list of questions on, “Have you noticed these things?” Chelle: (27:53) They asked you? TI005: Yes. Yes. I don’t know, I never knew that that was part of Parkinson’s.” |
| 5d. KNOW_ONLINE | Information from internet | TI004: “If it’s really, really relevant then I like the physical pages but there’s so much out there and so many different things in Parkinson's that aren’t relevant to (my husband’s) case that I usually start out on a website.”  TI005: “When he first laid it on me, she was at the hospital and I did what a lot of people do. I turned to doctor Google. I looked on Google and saw what it was, and I was like, “Oh. Oh. That’s what that is, okay.”” |
| 5e. KNOW_PAPER | Information from brochure, pamphlet, handout, book, etc | TI004: “Where did you first learn about PD psychosis? TI004: Probably in the book Parkinson’s For Dummies.” |
| 5f. KNOW_PRESENT | Information from presentations | TI001: “We might say, we might run a list, we get summaries of different workshops or presentations. I’ll read the presentations and then sometimes I say, “That’s really interesting.””  TI007: “Yes, I got more information by actually having the pharmaceutical company come and speak to... Parkinson’s Foundation was really good about bringing the pharmaceutical company in to speak to us when we were there where we could really ask the questions and find out more from them.” |
| 5g. KNOW_PROF | Information due to profession | TI007: “It’s probably more on my radar just because of the advocacy work that I do. I was part of the group that actually helped when they were coming out with Nuplazid and there was such an uproar over that medication when they did their first commercial. I’m on an advisory council for Parkinson's Foundation and we actually met with the pharmaceutical company and talked about why people reacted the way they did to that commercial.” |
| 5h. KNOW_TV | Information from television | TI002: “Well, one of the reasons why we’re here is because (my husband is) having hallucinations and I think we saw this thing on TV that said that there's a new medicine that might deal with that, and maybe he should get on it.”” |
| **6. MEDICATION** | Discussion of medication for PD/PD psychosis. |  |
| 6a. MED_INTERACT | Caregiver mention of psychosis med interacting with existing medications | TI001: But when you take eight or 10 medications or more, how do you really know what’s interacting, what isn’t?”  TI004: “It all depends on the side effects and whether they all play well together. He’s on so many different kinds of meds right now. He’s on Parkinson's drugs, he has heart issues and he’s taking all kinds of drugs for that. We’re walking a real fine line with the way all these are balanced together.” |
| 6b. MED_NOWANTMED | Caregiver replies that they don’t want their loved one on meds for psychosis, or do not feel like they need to be yet. | TI001: “Chelle: (51:46) Do you want him to? Or no? TI001: No. No, I don’t think it’s at a level to be.”  TI009: “Do you want him to be on a medication for the psychosis symptoms or not? TI009: Not really, just because I've heard that that can make other symptoms so much worse.” |
| 6c. MED_REASONWORSE | Caregiver’s reason for what would need to happen for them to advocate for medication for loved one’s PD psychosis. | TI004: “I would think if he spent more time having delusions and hallucinations and if the paranoia got a lot worse.”  TI006: “She would have had to been a danger to herself or to other people.”  TI007: “The symptoms would have to be progressing to the point where I was seeing them more often or that he was actually believing things.” |
| 6d. MED_TAKENO | Caregiver’s loved one does not take medication for psychosis symptoms | TI001, TI004, TI005, TI006, TI007, TI009 |
| 6e. MED_TAKEYES | Caregiver’s loved on does take medication for psychosis symptoms | TI002, TI003, TI008 |
| **7. STRATEGIES** | Strategies used by caregiver(s) to care for family member with Parkinson’s disease/PD psychosis symptoms. | TI001: “I have a neighborhood emergency team in place and it works.”  TI002: we wanted to get with a Parkinson’s specialist who would be on the cutting edge of research so that (my husband) could possibly participate in the clinical trials.”  TI003: “I could see the symptoms coming, so to speak. I was there, and I said, “You know, let’s breathe.” We’ve learned that. Gentle touch. Then I read to him, and that just calms him down.”  TI005: “Be nice. Don’t aggravate an already tenuous situation. You have this person who’s seeing things, who believes people are out to get her. Don’t aggravate it by saying, “Yes, they’re really out to get you.” Just talk it out.”  TI006: “like I said it took me back to parenting little toddlers and to diffuse situations, acknowledge, emphasize, distract. (Laughter.) That's the stuff that was the most helpful that gave me positive ways specific do's and don'ts of like I said, don’t try to convince her it's not there cause that's not going to work.”  TI007: “Exercise does help. If he is agitated sometimes going for a walk, encouraging him to talk.”  TI008: “[The neuropsychologist] will, quite often, give us some very concrete suggestions and type them up. We bring them home and we put them in a little book for everybody to read. They sign and initial how they read it.”  TI009: “I guess talking about it seems to help him quite a bit. When he says, “I just thought (our daughter) was in the room.” Then we talk about it and realize that it's not real. That helps it to go away.” |
| **8. TRIGGERS** | Triggers or circumstances that lead to increased instances of psychosis symptoms or episodes. |  |
| 8a. TRIGGERS_ENVIRON | Triggers related to time of day, location, environment. | TI002: “I think it's familiar surroundings and expectations of what should be around or whatever. That may be something that triggers the fact that he thinks the boys are here when they’re not.”  TI003: “He would, at a certain time of day, get very, very nervous. I could see the symptoms coming, so to speak.”  TI004: “The time of day. A lot of times he’ll be a lot worse in the evenings.” |
| 8b. TRIGGERS_LOWMED | Triggers related to low point of psychosis meds | TI001: “I think when he’s tired or his medication’s low.”  TI009: “He has his off periods. You know what off periods are. It's definitely worse when he is in those, with everything. With paranoia, yes. With everything, when he has an off period. It's all worse with that.” |
| 8c. TRIGGERS_MOOD | Triggers related to moods or emotions | TI001: “I think it comes along with either troubling memories or worries about something that might be going to happen or something.”  TI006: “It was making worse when you did what my father did with her and tried to convince her it wasn't happening, because then she'd get very upset about it.” |
| 8d. TRIGGERS_NOISE | Triggers related to loud noises, lights, machines | [TI003 only]  “The CNA came in, happened to be a man, deep voice, loud voice and it triggered a psychotic. Again the similar kind of episode that it’s hard to describe.”  “and it was the ultrasound and the machine and I wasn’t in the room. There was a curtain and I could see his face becoming more and more agitated and once again it related to machines.” |
| 8e. TRIGGERS_OTHER | Triggers that didn’t fit in another category | TI004: “If he’s having a bad day physically he will also have a bad day mentally. If he’s having a day where he’s freezing a lot, can’t go down stairs, is having trouble rising up out of the chair then that will be a day that he’ll be having trouble, or more hallucinations, more delusions.”  TI005: “Yes, I guess when she starts talking about how the job is out to get her, it’s because she’s been looking at something on Facebook and probably looking at coworkers’ posts.” |
| 8f. TRIGGERS_TIRED | Triggers due to being tired | TI001: “are there any triggers that you’ve noticed that either bring it on or make it worse? TI001: I think when he’s tired”  TI002: “It’s much more likely to happen when he’s tired, so around 6:00 on is when they happen. “ |
| 8g. TRIGGERS_UNEX | Caregiver describes that they are unable to predict when or why psychosis symptoms are triggered | TI003: “they were doing all these, trying to figure out what was happening and he for whatever reason, went into a full-blown psychotic, wild, at least verbally, violent episode”  TI005: “What are your clues that you get that it’s coming on or about to happen, if any? TI005: Nothing. She’ll just start talking about it.”  TI007: “Are there any clues or cues that you get that something like this is going to happen? TI007: No, not really.” |
